## Supplementary Materials for "Estimation of Respiratory Syncytial Virus-attributable Hospitalizations among Older Adults in Japan between 2015 and 2018: an Administrative Health Claims Database Analysis"

### SUPPLEMENTARY TABLES

**Supplementary Table 1. Definition of hospitalization main and secondary outcomes**

| Outcome | ICD-10 code† |
|---|---|
| **Cardiorespiratory diseases** | J00-J99, I21, I48, I49, I50, I63, I64 |
| **Respiratory diseases** | J00-J99 |
| **Influenza or pneumonia** | J09-18 |
| **Chronic lower respiratory disease** | J40-47 |
| **Chronic heart failure exacerbation** | I42-43; I50; I51.7 |

† International Statistical Classification of Diseases and Related Health Problems 10th Revision (ICD-10)-WHO, Version 2015. https://icd.who.int/browse10/2015/en#/ Note: In Japan, ICD-10 coding has been in use since 1990.

**Supplementary Table 2. Annual number of hospitalizations due to cardiorespiratory diseases (based on the source data [non-extrapolated]) by age group, January 2015-June 2019, Japan**

| Year | 60-79 years | ≥80 years |
|---|---|---|
| **Cardiorespiratory diseases** | | |
| 2015 | 84,732 | 97,227 |
| 2016 | 99,817 | 116,565 |
| 2017 | 117,904 | 141,718 |
| 2018 | 132,603 | 159,883 |
| 2019† | 74,344 | 91,664 |
| **Respiratory diseases** | | |
| 2015 | 42,021 | 55,466 |
| 2016 | 49,495 | 65,160 |
| 2017 | 56,546 | 78,874 |
| 2018 | 63,074 | 88,252 |
| 2019† | 35,718 | 50,524 |
| **Influenza or pneumonia** | | |
| 2015 | 17,107 | 23,897 |
| 2016 | 20,771 | 28,583 |
| 2017 | 23,509 | 35,546 |
| 2018 | 27,092 | 40,858 |
| 2019† | 16,086 | 24,983 |
| **Chronic lower respiratory disease** | | |
| 2015 | 4,244 | 3,593 |
| 2016 | 5,001 | 4,312 |
| 2017 | 6,092 | 5,296 |
| 2018 | 6,604 | 5,902 |
| 2019† | 3,742 | 3,317 |

| Year | 60-79 years | ≥80 years |
|---|---|---|
| **Chronic heart failure exacerbation** | | |
| 2015 | 14,041 | 22,830 |
| 2016 | 15,728 | 28,397 |
| 2017 | 19,591 | 35,789 |
| 2018 | 21,997 | 40,620 |
| 2019† | 12,741 | 24,184 |

† The year 2019 has incomplete data (until 30 June 2019)

**Supplementary Table 3. Annual number of RSV and influenza proxy DPC (Diagnosis Procedure Combination) hospitalizations (based on the source data [non-projected]), January 2015-June 2019, Japan**

| Year | Number of RSV proxy cases | Number of influenza proxy cases |
|---|---|---|
| 2015 | 35,288 | 6,928 |
| 2016 | 33,008 | 8,064 |
| 2017 | 46,452 | 14,388 |
| 2018 | 45,652 | 17,780 |
| 2019† | 13,992 | 17,160 |

† The year 2019 has incomplete data (until 30 June 2019)

### SUPPLEMENTARY FIGURES

**Supplementary Figure 1. Weekly number of RSV and influenza proxy DPC hospitalizations (based on the source data [non-projected]), January 2015-June 2019, Japan**

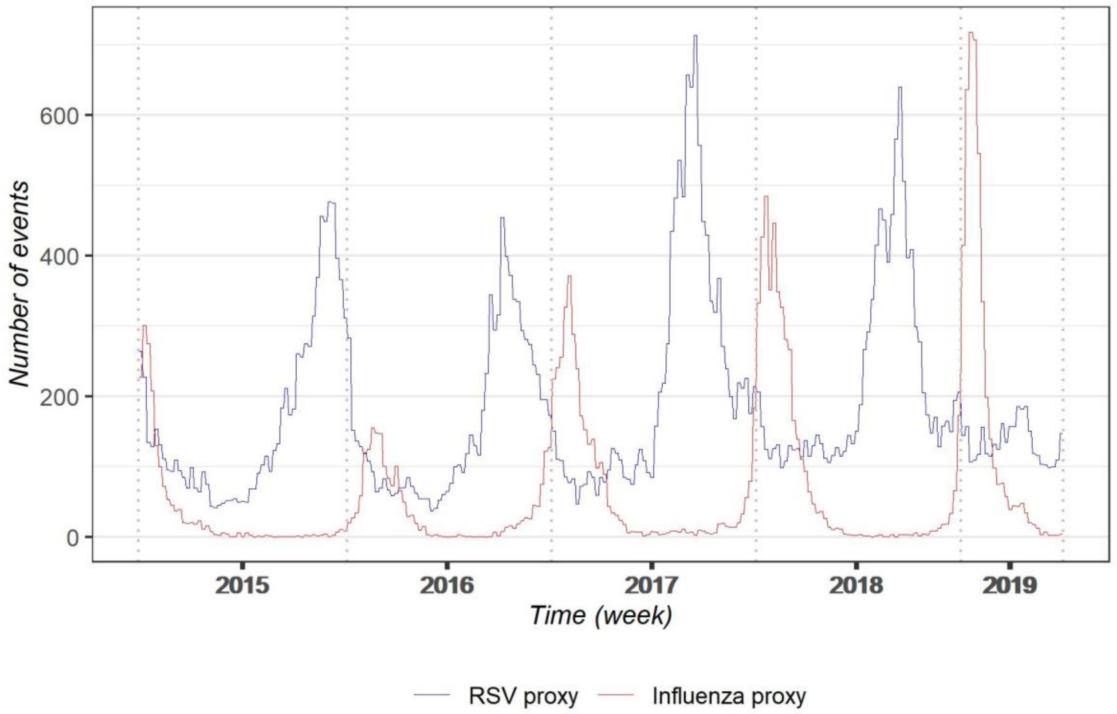
